# The Use of Behavioral Observation in Global Health: A Case Study from Rural Ethiopia

**DOI:** 10.1101/2025.05.15.25326691

**Authors:** Crystal M. Slanzi, Yuke Wang, Yang Yang, Elizabeth Schieber, Ibsa A. Ahmed, Abulmuen M. Ibrahim, Song Liang, Arie H. Havelaar, Sarah L. McKune

## Abstract

Behavior plays is critical role in global health research and practice. Indirect measures of behavior, such as surveys, are easier to implement and require fewer resources, and therefore are often favored over more direct measures, such as behavioral observation. Despite these advantages, indirect measures can also produce unreliable data. Direct measures, such as behavioral observation, produce much more reliable data, yet are often underutilized in global health. Moreover, when direct measures are used the descriptions of the methods related to data collection are often minimal making it difficult for others to replicate what was done. This paper is a case study outlining the data collection methods used in an epidemiological study that evaluated the pathways of infants’ exposure to *Campylobacter* spp. in rural Ethiopia. Included are details on the selection of outcome or target behaviors, the data collection software, the training of enumerators, and quality control measures as well as samples of the data produced through these data collection methods.

## 1. Introduction

It is widely recognized that human behavior plays a critical role in global health; examples include hygiene practices (Ngure et al., 2014), medication adherence (Migowa et al., 2009), and mosquito net use (Moon et al., 2016). To conduct assessments, develop interventions, and evaluate changes in various health outcomes, global health researchers and practitioners often seek to measure human behaviors, as both exposures and outcomes. For example, mosquito net use may be measured as an exposure protective against malaria infection; or it may be measured as an outcome, where prevalence of mosquito net use indicates the success or failure and sustainability of a malaria prevention program. The measurement of human behavior is thus critical in its provision of data that contributes to the investigation, surveillance, or assessment of myriad global health outcomes.

Such measurements are generally categorized as either direct (e.g., by observation and video recording) or indirect (e.g., by self-report through surveys and questionnaires, interviews, diaries, and focus group discussions). Although direct measurements are typically considered more objective and reliable than indirect measurements (Faukhauser, 2018), they often require more resources in the form of time, money, and overall effort, especially when data collection targets a large number of individuals (Madan et al., 2012; Mash & Foster, 2001) – a common characteristic of global health research studies. As a result, indirect methods are more commonly selected over direct observation, at the potential cost of lower quality data and less methodological rigor (Subramanian et al., 2009).

Indirect measures might be more frequently used due to advantages inherent in asking people to report how they behave rather than trying to directly observe what they do. One such advantage is that data collection methods that rely on reported rather than observed data are not dependent on the occurrence of the behavior and thus can be collected at any time the person reporting the data is available, allowing more flexibility in scheduling. Another advantage is that this approach requires less time, which not only saves time and money for the researchers and practitioners, but typically for the participants; this may make it more likely that the targeted population will agree to participate and provide information. Finally, these indirect methods also allow researchers to capture data on behaviors that are difficult, or sometimes impossible, to observe due to various reasons, including privacy concerns (e.g., sexual behavior, open defecation), extremely low frequency (e.g., occurs only one time per year), unpredictable occurrences (e.g., flooding related behavior), or the time of day (e.g., during the night).

Despite these noted advantages, it is important to recognize that the use of indirect methods can lead to biased data and subsequently false assumptions about what behaviors are occurring, as people are notoriously unskilled at reporting their own behavior (Althubaiti, 2016; Chan, 2009; Prince et al., 2008). One reason for this is that people tend to seek to portray themselves in a positive light (i.e., the social-desirability bias) by over- or under-reporting the frequency or duration of their own behavior (Krumpal, 2013). Over- or underreporting can also occur when participants misunderstand the questions being presented and report inaccurate information unintentionally (Rosenman et al., 2011). Reporting errors also occur due to recall bias, because people are simply unable to remember their own behaviors, even within a 24 hour time frame (e.g., Brusco & Watts, 2015). Individuals may also change their responses on later questionnaires following training, known as response-shift bias (Howard, 1980). Finally, being able to describe how to do something does not always translate into being able to perform the skill (e.g., Irehovbude & Okoye, 2020).

Due to the limitations of indirect measures, there have been calls to increase the use of behavioral observation and direct measurement in global health research and practice (Harvey, 2018; Weston et al., 2018; World Health Organization, 2009). A published summary of discrepancies between self-report data and observational data observed during a formative research study highlights just some of the issues that might arise with indirect measurement (see Harvey, 2018). For example, participants in a study evaluating mosquito net usage reported that they remained in their nets for the duration of the night; however, observers noted that the participants failed to report that they frequently exit and re-enter their nets throughout the night, often failing to close it correctly to prevent mosquitos from entering (Harvey et al., 2017). The need for increased monitoring and direct measure has also been noted in water, sanitation, and hygiene (WASH) research (e.g., Dey et al., 2019); a bid echoed by the World Health Organization’s (WHO) that has stated that “direct observation is the gold standard to monitor compliance with optimal hand hygiene” (WHO, 2009, p.103).

Behavioral data have also been underutilized in epidemiological modeling (e.g., Funk et al., 2015; Funk et al., 2010; Weston et al., 2018). For example, some early models of the COVID-19 pandemic used policy changes, a fixed parameter, without the inclusion of factors such as adherence to those policies and the compliance fatigue that would set in over time (Rahmandad et al., 2021). When behavioral data are included in models, they are most often based off indirect measures such as self-report surveys that include questions about beliefs and intent. A recent review of statistical methods in infectious disease modeling found that only 16 of 42 papers included any behavioral data, and none of them contained data from direct measurement (Weston et al., 2018).

In recent decades, some researchers have incorporated behavioral observation into quantitative exposure assessment (e.g., Teunis et al., 2016). One example of this is an exposure assessment, referred to as SaniPath, which has been primarily used to quantitatively assess the exposure to fecal contamination indicated by *Escherichia coli* through various pathways (Robb et al., 2017; Wang et al., 2017). In several of the SaniPath studies (e.g., Robb et al., 2017; Wang et al., 2017), enumerators recorded the location, duration, and frequency of activities of their participants; however, frequency events were recorded as an overall number (e.g., 10 times per hour) and not in the order in which the events or activities occurred. In addition, in the SaniPath papers that incorporated direct measures, (e.g., Robb et al., 2017; Wang et al., 2017), many, if not all, specify that the observations were conducted by “trained” enumerators, but did not include training methods, such as how it was determined that the amount of training was sufficient to ensure they were able to record reliable data.

Omitting details on how enumerators are trained is not uncommon in articles on global health topics. Even those that call for an increase in the use of direct measures (e.g., behavioral observation) have left out descriptions of how enumerators were trained to competency (e.g., Harvey, 2018). Moreover, when papers in global health include behavioral data (e.g., Harvey et al., 2003; Teunis et al., 2016), detailed descriptions of the methods related to data collection, such as the operational definitions of the behaviors being recorded, data quality control measures, and details about the recording modality (e.g., pen and paper or electronic application) are also not typically included. Although this is likely due to these details being out of the scope of these research articles, the information is not outlined elsewhere, which limits a reader’s ability to gauge the validity of the data presented or replicate the methods.

Given the limited use of direct measurement (e.g., behavioral observation) among reported methods in global health literature and the lack of descriptions of the methods related to data collection, the purpose of this paper is to provide an example of how behavioral observation can be effectively implemented in global health research using a case study from rural Ethiopia.

## 2.0. Case study: EXCAM

In 2018 a team of researchers from the University of Florida, Emory University, and Haramaya University designed a study to examine the pathways of infants’ exposure to *Campylobacter* spp. using methodological advances that supported improved quantification of exposure. The study, entitled *Exposure Assessment of Campylobacter Infections in Rural Ethiopia* (EXCAM), was a prospective observational study of infants in rural Haramaya, Ethiopia. The study aimed to 1) characterize children’s space-time behavioral patterns in their living environment (e.g. contact/contamination interfaces within the household and neighborhood); 2) measure *Campylobacter* spp. across the space-time contact/contamination interfaces and quantify them using qPCR and, in children and mothers/caregivers, 16S rRNA sequencing techniques; and 3) assess children’s total exposure and the contributions of exposure pathways to infection and disease risks. To achieve the first aim, local enumerators conducted direct observation of 79 children between 4-8 months of age and 76 of those same children at a second timepoint when they were between 11-15 months of age. Children were recruited from the *Campylobacter Genomics and Environmental Enteric Dysfunction* (CAGED) study (Havelaar et al., 2022), which aimed to identify the prevalence, diversity, and reservoirs of *Campylobacter* infections in infants.

## 3.0 Methods

### 3.1 Setting

EXCAM took place in 12 kebeles in rural Haramaya, Ethiopia. This area of Eastern Ethiopia is highly dependent on agriculture, with most families relying primarily on production of khat, a cash crop, for income. Communities are predominantly Muslim, and family centered; have low education levels, minimal infrastructure, and poor hygiene and sanitation; and often have a mix of livestock and crops. Multiple family members may have households that are physically adjacent and connect to form a homestead. Each household is typically one building, in which children, adults, and often livestock sleep. Outside, and sometimes shared by households within a homestead, there is a designated cooking area and often a stable/shed for livestock or other assets. Most households do not have access to or use a latrine nor a reliable source of safe drinking water.

### 3.2 Study Population

The study population of mothers with newborns was recruited directly from the CAGED study (Havelaar et al., 2022). At the time of enrollment in CAGED, mothers were asked if they would be willing to participate in the second study. If they were willing, study team members provided information and took them through an informed consent process for EXCAM. A total of 79 women and children participated; 79 of these children were observed once and 76 twice.

### 3.3 Outcome Variables or Target Behaviors

We went through several steps to identify and define the behaviors that local enumerators would take data on for the study. The selection of target behaviors was initially guided by the states, which included compartments or the infant’s location (e.g., carried on mom’s back) and activities (e.g., sleeping) used in prior literature on exposure assessment (e.g., SaniPath; Wang et al., 2017), as well from observations conducted during formative research (Chen et al., 2021). We determined which behaviors may be most relevant based on typical caregiver and infant behavior in the region. Once the target behaviors were chosen, we piloted data collection in the field to identify any transmission related behaviors that were not on our initial list and remove ones that were irrelevant. Beginning in February 2022, the enumerators began making a note in Countee at the end of each session indicating if the infant had started crawling or walking.

Based on those observations we also developed operational definitions for each of the behavior or activities and determined whether they were to be measured as frequency-based or duration-based event. For example, pica was a frequency-based measure defined as any non-food items entering the mouth completely where the mouth closes around the item and swallowing is possible (see Table 1 for a complete list of target behaviors and operational definitions). To ensure that our operational definitions were sufficiently clear and that all trained enumerators were able to identify the same behavior when they observed it based on the definition, we conducted trial observations in the field. Specifically, two different data collectors independently observed the same infant. We then assessed the level of agreement between the data collected from each enumerator. Definitions were finalized following discussions of disagreements and further field testing.

**Table 1.** Summary of Recorded Events, Subcategories, Dimension, and Operational Definitions.

| Recorded Events and Subcategories | Dimension | Operational Definition |
| --- | --- | --- |
| Location |  |  |
| On homestead | Duration | The infant is somewhere within the homestead. This may include structures surrounding the home such as a stable, kitchen, or grass around the house. |
| Off homestead | Duration | The infant has left the homestead. For example, going to the market or being carried into a field of crops. |
| Compartment |  |  |
| Carried mom arms | Duration | The infant is being held in the arms or lap of their mother and is off the ground |
| Carried mom back | Duration | The infant is swaddled onto their mother's back |
| Carried other arms | Duration | The infant is being held in the arms or lap of an adult that is not their mother and is off the ground |
| Carried other back | Duration | The infant is swaddled onto an adult's back that is not their mother |
| Down with barrier | Duration | The infant is laying down without the assistance of a person, but with some type of barrier between the child and the ground. For example, a mat or blanket |
| Down bare ground | Duration | The infant is laying down without the assistance of a person, but with some NO barrier between the child and the ground. |
| Activity |  |  |
| Sleeping | Duration | The infant has had their eyes closed and suspended movement for at least 1 minute. *note: do not end "sleep" unless the baby has had their eyes open for 1 minute, or if they cry or move into one of the activities below. Thus, if the baby opens their eyes and adjusts its limbs then closes their eyes and settles in, keep "sleep" running instead of changing the activity |
| Awake | Duration | The infant shows signs of being awake without being engaged in another activity. For example, their eyes are open, and they are moving around, they are cooing and reacting to others' attention, light, etc. IF they were previously sleeping, they must show signs of being awake for 1 minute. |
| Bathing | Duration | The infant is being washed with water or a wet cloth |
| Drinking breastmilk | Duration | The infant is suckling milk from a breast. If they release the breast and return within 5s, keep this activity going. If the baby switched to the other breast, stop and start this activity again |
| Drinking water | Duration | The infant is consuming water. If they stop drinking and return within 5 seconds, keep this activity going |
| Drinking other | Duration | The infant is consuming any other liquid (for example: animal milk). If they stop drinking and return within 5 seconds, keep this activity going |

**Table 1 Continued**
| Recorded Events and Subcategories | Dimension | Operational Definition |
| --- | --- | --- |
| Activity: Eating<br>Injera<br>Raw plant<br>Other | Frequency | Press this button for every bite taken then indicate what type of food was consumed (injera, a raw plant, or other). The activity key that should be running during periods eating is “awake” |
| Activity: Mouthing<br>Baby hand<br>Mom hand<br>Other adult hand<br>Child hand<br>Fomite | Frequency | Press this button any time the baby has one of the following pass the plane of their lips but is not eaten. Then define what the item is: the baby’s hand, their mother’s hand, any other adult’s hand, a child’s hand, or a fomite. Note: count a single event if the hand or item remains in contact and a new event it makes contact with the mouth ends and starts again. |
| Activity: Touching<br>Mom hand<br>Other adult hand<br>Child hand<br>Fomite<br>Livestock | Frequency | Press this key any time the baby’s skin (not clothing) touches one of the following items then define what item touched the baby. This can be initiated by the baby or another person. The items are: mother’s hand, another adult’s hand, a child’s hand, any fomite, or livestock. |
| Activity: Pica<br>Feces<br>Soil<br>Other | Frequency | Press this button any time any of the following items is placed into the mouth such that it could be swallowed: feces, soil, or other.<br>*note, mouthing a fomite is not necessarily pica if it cannot be swallowed, for example a cloth or large object. But if it can be swallowed and enters the mouth, it is considered pica, even if someone intervenes to block swallowing For example, putting a rock in the mouth. |

*Note.* The subcategories (qualifier keys) for location, compartment, and activities have different operational definitions, but the subcategories for eating, mouthing, touching, and pica do not.

**Table 2.** Summary of Behavioral Observation Data.

|  | Timepoint One | Timepoint Two |
| --- | --- | --- |
| Number of Infants Observed | 79 | 76 |
| Mean age in weeks (range) | 26 (18- 37) | 53 (48-66) |
| Hours of observation | 702 | 608 |
| Location – average % of time on homestead | 99.6% | 97.8% |
| Location – average % of time off homestead | 0.4% | 2.2% |
| Compartment – average % of time carried by mom | 40.3% | 27% |
| Compartment – average % of time carried by other | 14.4% | 6.9% |
| Compartment – average % of time down with barrier | 45.1% | 49.8% |
| Compartment – average % of time on bare ground | 0.2% | 16.4% |
| Activity – average % of time awake | 60.9% | 76.% |
| Activity – average % of time sleeping | 21.9% | 12.1% |
| Activity – average % of time drinking | 16.8% | 11.7% |
| Activity – average % of time bathing | 0.3% | 0.3% |
| Eating – average rate all subcategories | .35/hr | 2.20/hr |
| Mouthing – average rate all subcategories | 13.19/hr | 10.69/hr |
| Touching – average rate all subcategories | 20.15/hr | 37.11/hr |
| Pica – average rate all subcategories | .005/hr | .67/hr |

We chose target behaviors across three categories which included both frequency- and duration-based behaviors or events: location (duration), compartments (duration), and activities (both duration and frequency; see Table 1). Locations included on- and off-homestead, compartments included being carried, or being down with or without a barrier, (i.e., protecting the infant from direct contact with bare soil) and activities included drinking (including breastmilk), sleeping or awake, and bathing; all of which were recorded as durations. All the sub-categories (e.g., down with barrier for compartment) were mutually exclusive, which meant that only one could be recorded at any given time. In addition, at least one location, compartment, and duration-based activity always had to be selected, so that the durations of all behaviors would account for 100% of the observation time. For example, under the category of activities the distribution of time for the behavior of the infant may have been as follows: drinking breastmilk (24%), sleeping (43%), awake (28%) and bathing (5%). Events were behaviors that were recorded as frequencies and included mouthing, being touched by others, touching others, and eating. On the Countee application, behaviors are recorded as they occur resulting in an output showing both the behaviors occurring at each second of the observations and the order in which they occur. This provides a level of precision in the measurement of transmission behaviors that, to our knowledge, is novel to quantitative exposure assessment.

### 3.4. Data Collection Technology: Countee

To record the frequency, duration, and sequence of behaviors during the observation the enumerators used an application called Countee© (Peic-Gavran & Hernández Eslava, 2020), which is free to download in multiple countries on both iOS and Android (see Slanzi & Fernand, 2024 for a tutorial on Countee). In addition to the main keys or icons on the home screen for each target behavior, the tablet or phone-based application allows subcategories or qualifier keys (e.g., if the main key is eating, the type of food could be the subcategory) to be added for each target behavior that appears as a drop-down menu during data collection (Figure 1). To record a behavior, the enumerators pushed a key or icon on the tablet screen and then select the appropriate subcategory from the drop-down menu. For frequency events, the key had to be touched a single time, and for duration events (e.g., breastfeeding) the key had to be touched a second time when the behavior ended. If an enumerator pressed a key in error, they could cancel it in the drop-down menu. If an enumerator did make an error of commission, they could press an error key immediately after the error. By doing so, team members who cleaned and analyzed the data could remove the entry immediately prior to the “error” entry in the output file. The data were saved in Dropbox as a CSV or Excel file and then were imported in R (R Core Team, 2023) for cleaning and analysis (see Figure 2 for a sample output). An additional benefit of Countee is that internet access is not required during data collection sessions; files are saved in the application and uploaded when enumerators were able to connect to the internet. This was essential for this project due to instability in both electricity and internet connectivity across rural communities and prevented data loss in regions where stable connections were unavailable.

**Figure 1.**
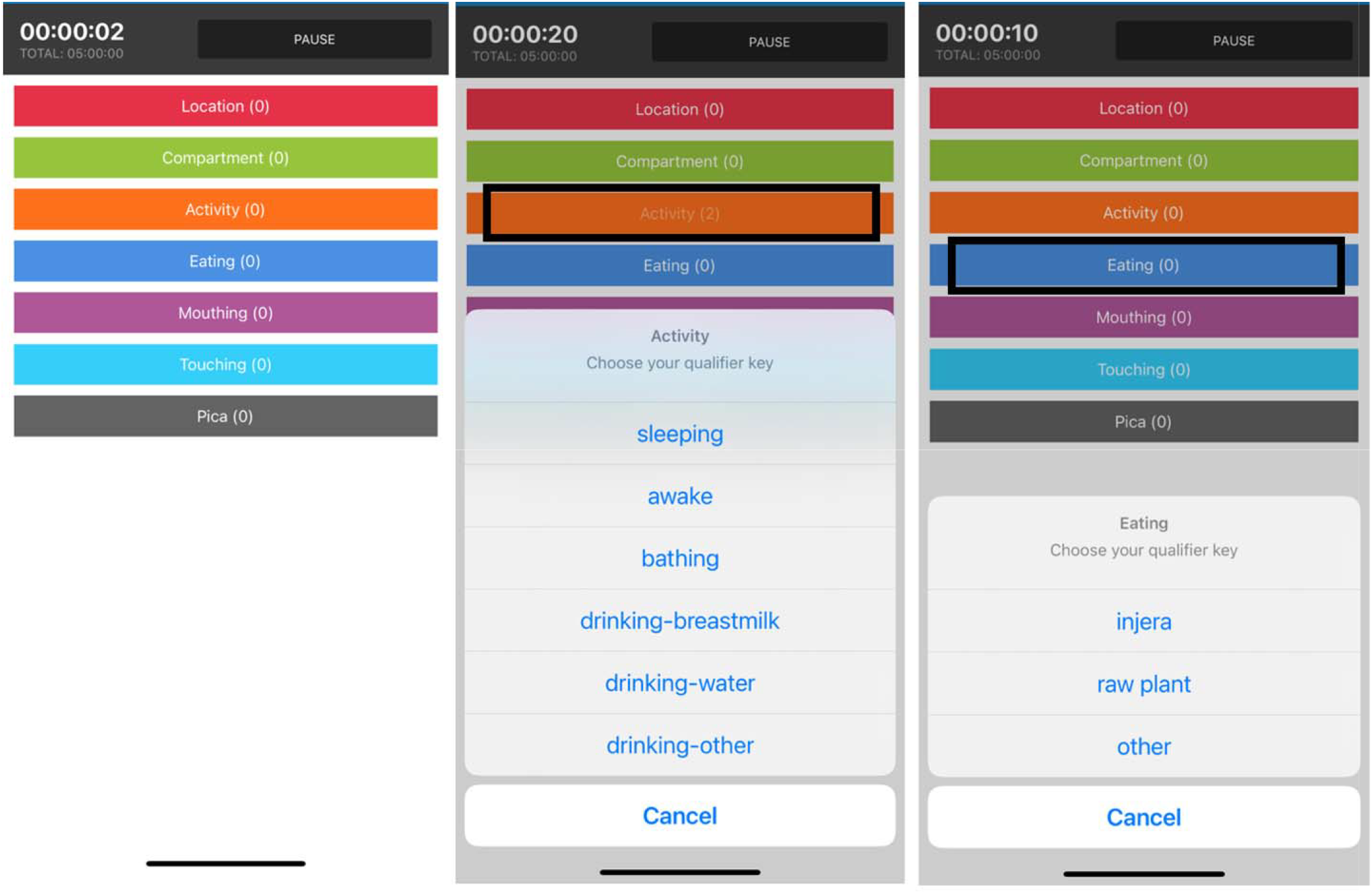
Samples of Countee Screens for Data Collection Including the Main Screen and Subcategories for Activity (Duration) and Eating (Frequency)

**Figure 2.**
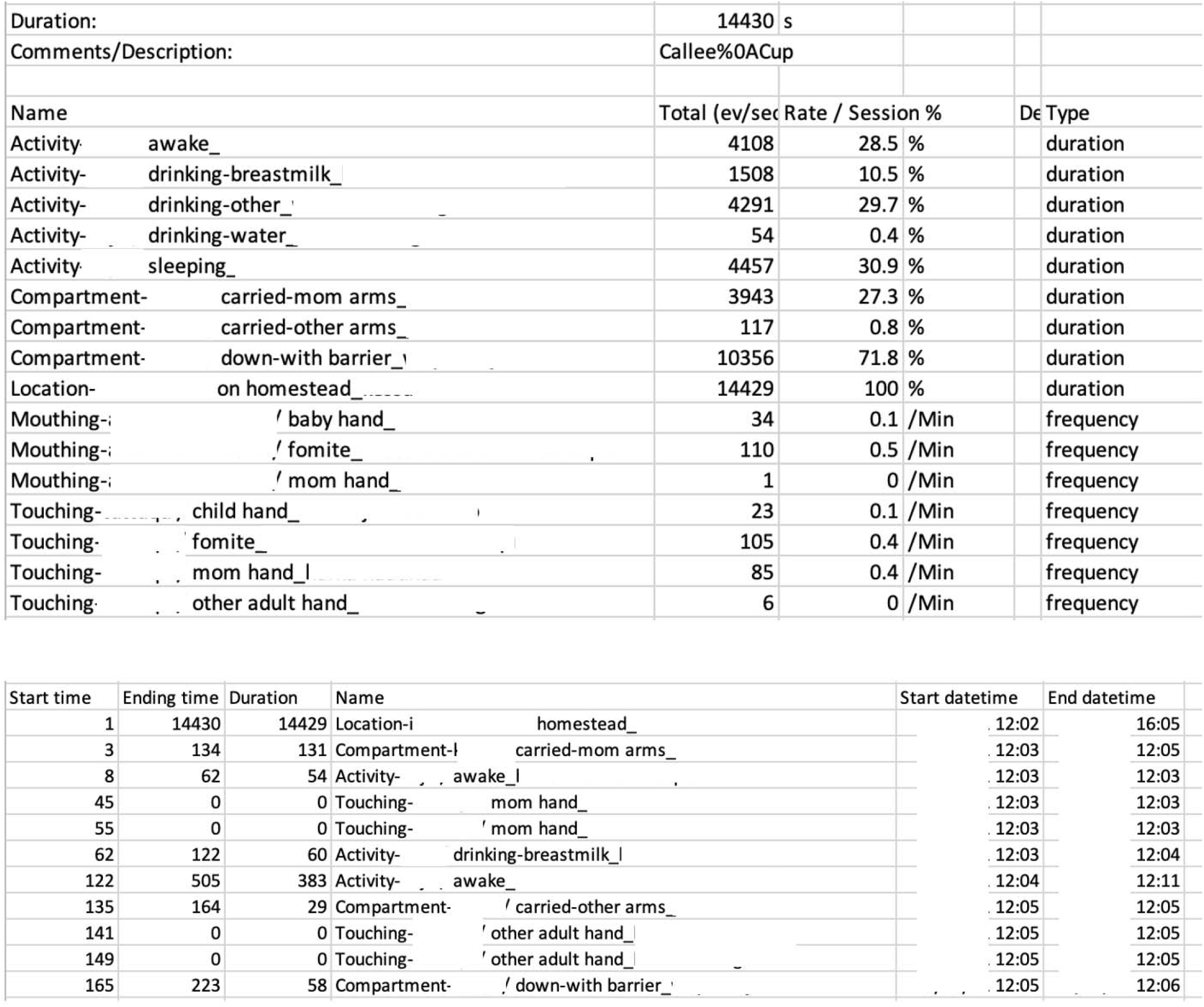
Sample of Summary and Real Time Data Output from an Observation Note. The top figure displays the summary of all events recorded during the observation as well as the observation, date, time, and duration. The bottom figure shows a sample of the second-by-second recording of events from the same observation.

### 3.5. Enumerator Description

Four female enumerators from the same community where data were being collected were hired for the duration of the study. Local women were intentionally hired as data collectors to increase the comfort level of the women being observed. Hiring local women who have a better understanding of local customs and cultures drew less attention from others living in the same kebele and was less intrusive in the life of a new mother than a male observer.

### 3.6 Training Enumerators

Prior to conducting any training on collecting data during a behavioral observation, the enumerators were taught how to set up and use Countee. Competency-based training (a.k.a., behavior skills training) was used to teach the enumerators to record data reliably during an observation. Competency-based training includes the following steps: 1) providing verbal and written instructions, 2) demonstrating how to collect the data either live or on a video, 3) allowing time for rehearsal or practice, and 4) providing feedback which continues until the trainee is able to demonstrate the target skill with specified level of accuracy on two to three separate occasions (Vladescu & Marano, 2021). For this study, enumerators were given a list of target behaviors with operational definitions for each and then shown what each behavior looked like on a video of an infant. They were then asked to record infant behaviors they observed in a series of videos using Countee. Most of the videos were of infants, none of whom were participants in the study, that were recorded with caregiver permission from the study the region was being conducted. Additional videos were also recorded of infants outside the region or were from YouTube (e.g., a YouTube video of a child eating solids and taking many bites as none of the local videos featured infants eating solids) and it was not possible to record more from the local region in that moment.

To determine competency, the data collected by the enumerators had to reach 80% agreement with a trained observer, a practice referred to as inter-observer agreement (Ostrov & Hart, 2013). We used a scored interval agreement method to calculate agreement during training videos for this study, and during practice observations. This was done by breaking the sessions into 1-minute intervals and comparing the frequency and duration of recorded events by two different enumerators in each interval, only intervals during which one or both observers indicated a behavior occurred (i.e., a scored interval) were compared. The percentages across all scored intervals for each target behavior are then averaged for an overall percentage. Scored interval agreement was calculated by hand by the trainer. Comparing data across intervals (e.g., data recorded each minute) is more precise than comparing it across an entire observation session because it prevents potential false rates of high agreement by ensuring that the behaviors are recorded around the same time, increasing the likelihood that the enumerators recorded the same behaviors. For example, if both enumerators record 15 occurrences of a target over the whole session, it is possible that they did not record the same 15 occurrences.

The videos used in practice sessions ranged in duration from 30 s to 10 mins and increased in complexity (i.e., included more target behaviors) until a final novel test video was provided. To be considered adequately trained, each enumerator had to record data from the test video three separate times and, as with other videos, it had to correspond with those taken by a trained enumerator with at least 80% agreement. If an enumerator was unable to record reliable data with the test video, they continued to practice and receive feedback using novel videos. Once they were ready (e.g., recording data with 80% agreement during test videos), another novel video was provided to test mastery. After achieving mastery with the videos, enumerators participated in two, approximately 1-hour observations in the natural environment with the trainer to test their skills.

### 3.6. Observation Schedule

Once training was complete, behavioral data for the quantitative exposure assessment were collected during two, approximately 10-hour observations for each participant approximately 6 months apart; the first at 4-8 months and the second from 11-15 months of age. The 10-hour observations were split into two, 5-hour sessions on the same day, each conducted by a different enumerator. The number of sessions and duration of each observation were chosen to ensure collection of sufficient behavioral data for the accuracy of the transmission pathway assessment. The session lengths were also designed to reduce reactivity (i.e., the Hawthorne Effect) of the mothers being observed, even though we expected it to be low already by using female enumerators local to the area (Chen et al., 2015) The sessions were split into two, 5-hour sessions, to reduce enumerator fatigue and errors, sometimes referred to as observer drift (Ostrov & Hart, 2013). Although, 5 hours is still a long time, the enumerators were able to take short breaks when the infant was sleeping, as no other behaviors would have been occurring at that time. Seasonal shifts in agricultural work were also taken into consideration when scheduling observations as they can alter daily routines, particularly for families growing khat or staple crops. During peak harvest weeks, caregivers are more likely to leave the homestead early in the morning, sometimes leaving infants with relatives for extended periods. Because of these fluctuations, we adapted data-collection schedules to align with typical breaks in fieldwork, ensuring enumerators could capture a broader range of childcare behaviors throughout the day.

### 3.7 Quality Control Methods

All data were uploaded directly from Countee into Dropbox by the enumerators at the end of each observation. To ensure that no data were missing, a record of which participants or households had been observed was created and cross-checked with what was in Dropbox. Each data file was opened, and calculations were conducted to assess potential errors in the data by evaluating if the duration or frequency of events seemed too high or too low given the age of the infant, averages across data samples, or in comparison to other events (e.g., time spent awake). For example, given that a location and activity (e.g., sleeping, awake) had to be selected for the entire duration of the 5-hour observation, those events should add up to 100% of the observation time (or very close to it). Finally, data were spot checked for incompatible events, such as behaviors recorded while the infant was sleeping, or no behaviors or very few behaviors recorded while the infant was awake. If potential errors were noted during spot checks, additional training was provided to the enumerator who made the error.

In addition to spot checks of the data for quality, on three separate occasions, enumerators observed the same infant at the same time, and the data they collected were compared for agreement. We used a proportional agreement method to calculate IOA automatically through the Countee application website (https://www.counteeapp.com/ioa/) to check agreement during the quality control checks. After uploading the Excel spreadsheets for each observer and indicating the interval duration (1 minute) the website produces a CSV file with the agreement percentages for each behavior (see Figure 3 for an example and Slanzi & Fernand, 2024 for a more thorough description of this feature).

**Figure 3.**
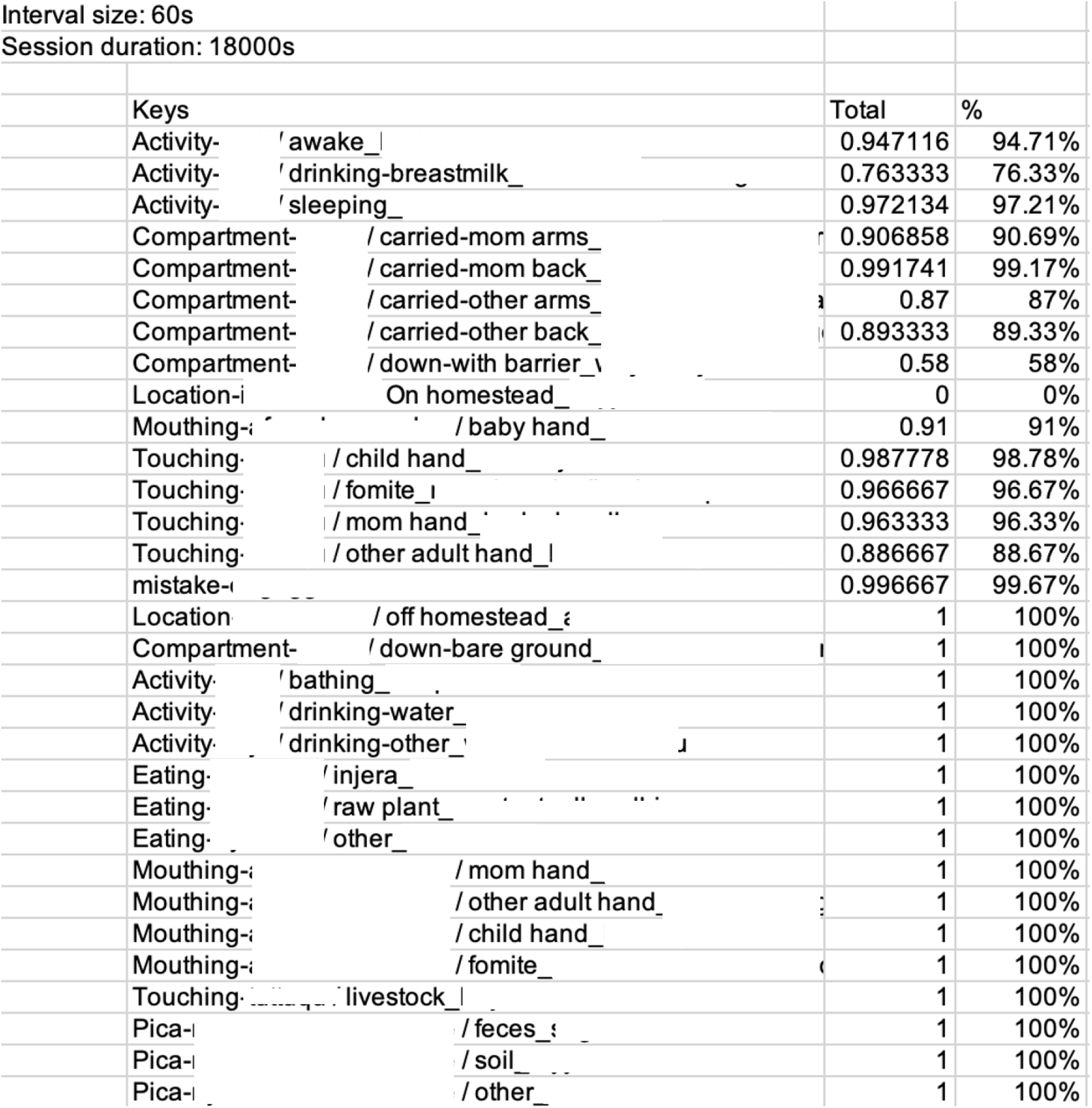
Sample of Agreement from One Quality Control Data Collection Session for Frequency Events Note: The criterion for agreement indicating reliable data was set at 80%. Based on this observation, for 29 of the 31 target behaviors, the criterion was met. For the two that were below (drinking breastmilk & down with barrier), which were both duration-based behaviors, booster training was provided. The “mistake” line refers to the key that enumerators pressed following an error of commission (pressed a key by accident).

The only difference between this method and the scored interval session mentioned above is that all intervals are compared, even ones in which no behaviors were recorded by either enumerator. This method was chosen over the scored interval method for these data due to the length of the observation sessions (i.e., 1 hour maximum in training vs. up to 5 hours for the study). This method decreases the potential for false low agreement that can occur for long observations when only scored data interval periods are included. For example, suppose in a 5-hour observation, the two enumerators each recorded the same behavior in one interval, only one recorded it in a second interval, and then no behaviors were recorded for none of the remaining intervals. The agreement using a scored interval would be 50% (one interval of agreement out of two scored intervals) as it would fail to account for the fact that agreement for the non-occurrence of behaviors was quite high for most of the session. This method was not required during training as there were few intervals without the occurrence of a behavior because the videos were short, and the in-person observations were planned during periods of higher activity when the infants were awake.

The data for the quality checks were done during regularly scheduled behavior observations, therefore we used the data collected by the enumerator that was assigned to that observation period in the study rather than the “extra” person that was overlapping them. During these overlapping sessions, the enumerators started their data collection session in the application at the same time, sat apart from each other, and did not speak during the observation. One of the reasons for doing this was to assess observer drift, or the tendency for enumerators to change the way they take data (e.g., including behaviors that are not part of the operational definition) which may result in errors. If the agreement between the enumerators dropped below 80%, additional booster training was provided.

## 4. Results

### 4.1. Training and Quality Control

Training continued for all the enumerators until they were able to reach 80% agreement with the data collected by the trainer in videos and in-vivo at an infant’s home (see Figures 4 for a sample of the training data). On three separate occasions, twice in September 2021 and once in March 2022, two of the enumerators observed the same participant at the same time. The data were then assessed to determine the degree to which they were recording the same behaviors at the same time and for the correct duration (see Figures 3 and 5 for a sample of rates of agreement). If the rates of agreement fell below 80% for a specific behavior due to observer drift (which they did) both enumerators participated in booster training. In the example in Figures 3 and 5, two of the 12 duration-based events fell below 80% agreement (i.e., drinking breastmilk, 76% and down with barrier, 58%), but none of the 17 frequency-based events did. Booster training was provided for each of these two behaviors.

**Figure 4.**
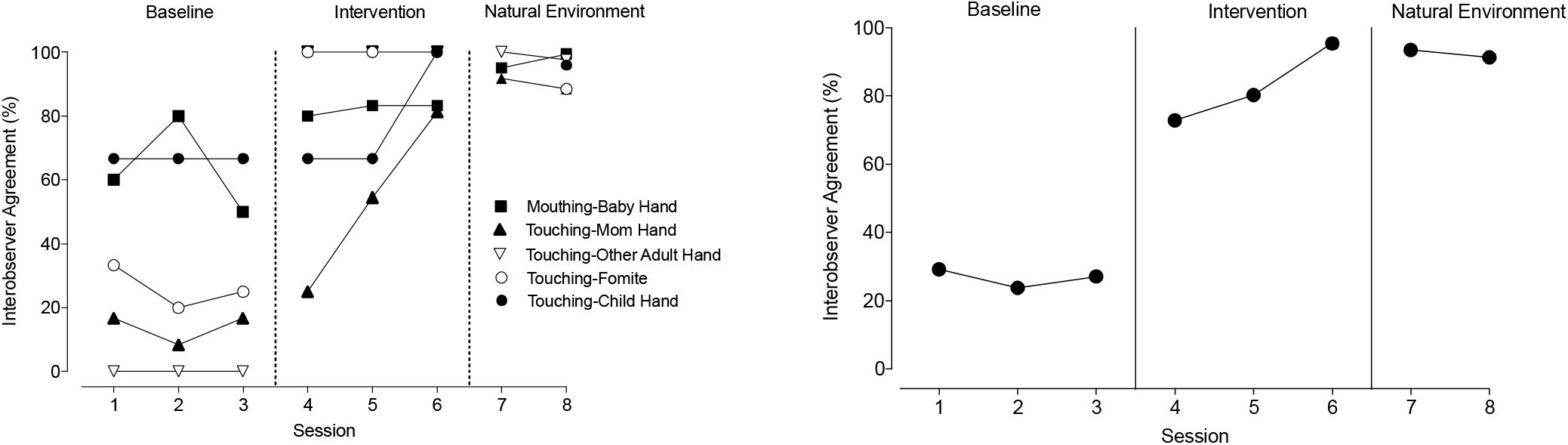
Graph Depicting a Sample of Percent Agreement of Frequency-Based Behaviors Between the Trainer andan Enumerator in Training at Baseline, During the Training (Intervention), and in an Infant’s Home. Note. The figure on the left depicts agreement for each target behavior individually and the figure on the right depicts agreement for all behaviors combined.

**Figure 5.**
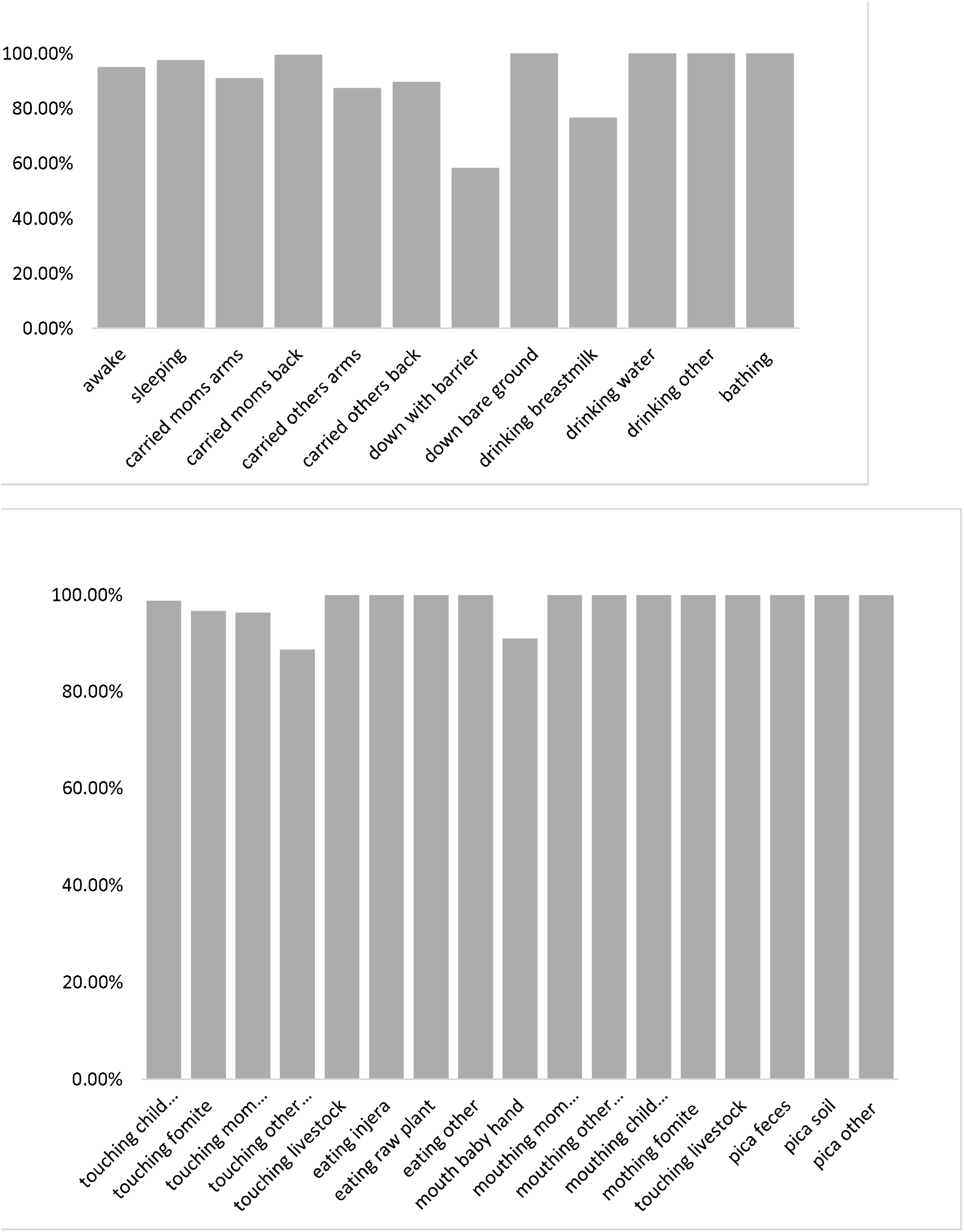
Proportion Agreement Rates for Duration-based Behaviors (top) and Frequency-based Behaviors (bottom) for One Spot Check for Reliability of Data Collection Note: the horizontal line represents the targeted agreement rate between two observers of 80%.

### 4.2. Behavior Summary

There were two time points for data collection. The first was from May 2021 to December 2021 and the second was from December 2021 to June 2022 (see Table 3 for a summary of behavioral data and Figure 6 for a sample heat map of one participant’s behavior). The average age of the infants in the first timepoint was 26 weeks (approximately 6 months) and at the second timepoint was 52 weeks (approximately 1 year old). Between these two timepoints drinking (which included breastfeeding) decreased by 5% of the total time and the rate of eating solid foods increased by almost two occurrences per hour (from .35 to 2.21). This reflects an increase in the infants eating solid foods and subsequently breastfeeding less often. For 47 of the 75 observations at the second timepoint, starting on February 3^rd^, 2022, enumerators began making a note in the Countee application (it is possible to type session notes in the application before ending the session) indicating if they observed the infants crawling or walking. Of these 47 infants, four (6%) of the infants were walking and the other 43 (94%) were crawling. Given that 100 % of the infants for whom these data were recorded were walking or crawling it is likely that many if not all the infants observed at the second timepoint were also walking or crawling even if the occurrence was not recorded. This behavior change likely led to the increases in the time infants spent being down on the bare ground (16% increase), the rate of pica (from .005/hr to .66/hr), and the rate of touching items or other people (from 20/hr to 37/hr) as the infants were able to independently reach and access more items and people in their environment.

**Figure 6.**
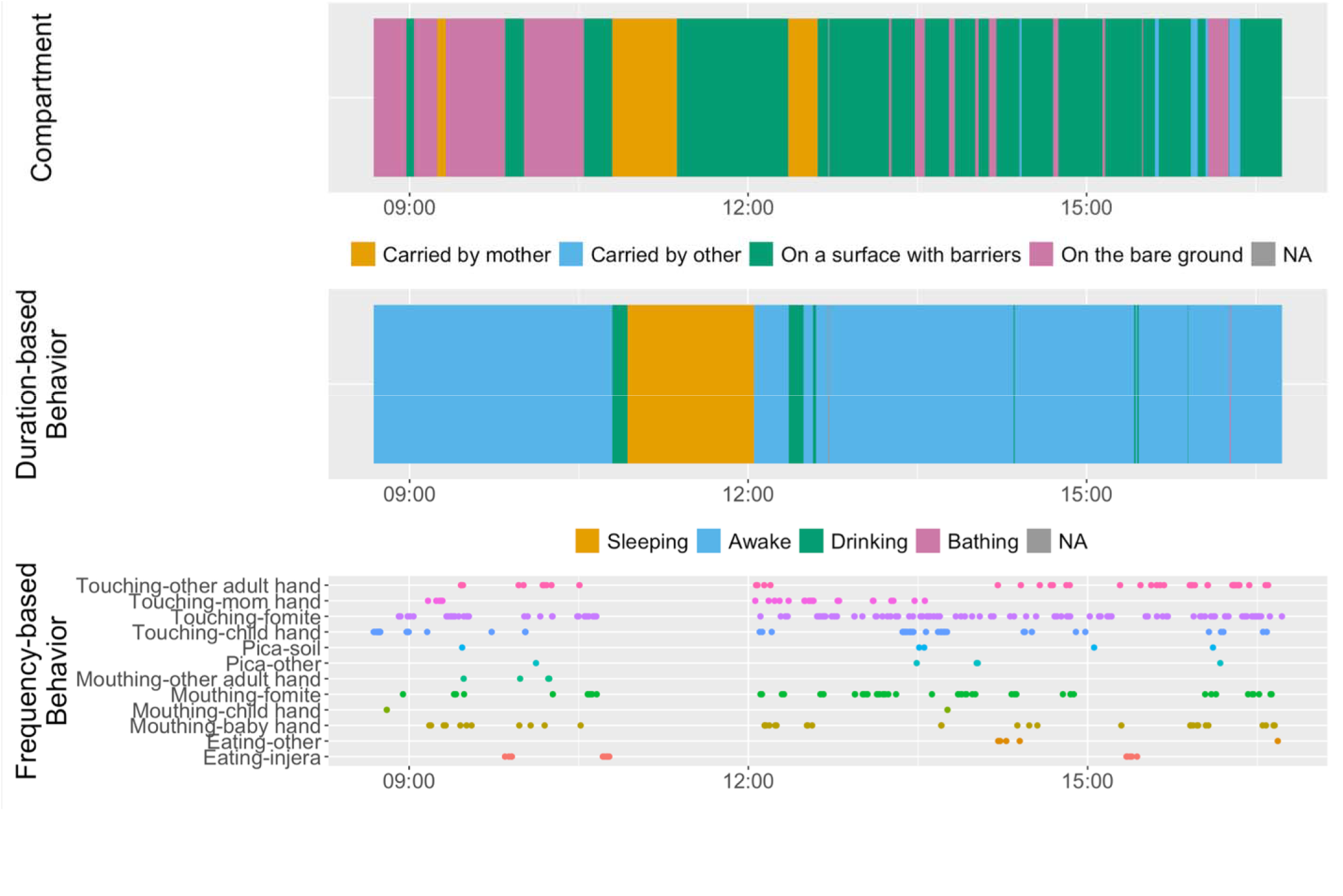
Sample heat map or piece of one.

## 5.0 Discussion

### 5.1 Implications for Research and Practice

The use of behavioral observation in this study led to an improvement in the accuracy of the exposure assessment. Specifically, the methods used to design the data collection system led to the collection of higher quality, more reliable data than self-reported methods would have yielded. One reason for this was the use of the Countee application, which allows for the recording of events and behaviors in sequence with timestamps. As previously mentioned, in series of studies using the SaniPath approach, data were either taken on paper or with an electronic application called Open Data Kit (ODK) – neither of which allowed for high-resolution timestamp recording (Teunis et al., 2016; personal communication with the SaniPath team). The use of Countee, with the programmed main keys and sub-category qualifiers, made it easy for the enumerators to record a high volume of data in a short time. Further, during pilot testing of the data collection systems, three local staff tested ODK and Countee during natural environment observations, and they unanimously preferred Countee. For example, the programmed subcategories allowed the enumerators to record the activity (e.g., drinking) and subcategory (e.g., water) in a matter of seconds by tapping the screen twice as opposed to having to scan a page, find the event, record an event, and then record the subcategory (potentially by writing it out), which would have been required with ODK. As a result, the enumerators recorded 23,677 and 30,372 frequency-based activities on the first and second observations (mean of 18 to 23 events/hr), respectively, in just over 1300 hours of observation of infants who slept for a considerable portion of each session. This is a high volume of data, especially considering that in a similar SaniPath study, enumerators recorded 1684 events in 500 hours (mean of 3 events/hr) when observing infants and children up to the age of five who would have been awake for longer durations and more active due to their age (Teunis et al., 2016). The level of training the researchers provided to the enumerators, which was based on skill acquisition and competency rather than time, likely contributed to increased quantity and detail of the collected data. Because other published studies that used behavioral observation did not include details regarding the training that was provided to enumerators or if they were required to meet determined competency levels prior to conducting data collection sessions, it is unknown the extent to which human errors of omission and commission have affected data quality in those previous studies.

Inclusion of direct measures of behavior in exposure assessments that characterize transmission of a given pathogen is likely to improve accuracy; and increased accuracy in these models may afford the scientific and practice communities’ new opportunities to improve the effectiveness of behavioral interventions aimed at reducing infection rates. Enteric infections, which are the focus of this case study, lead to 1.7 billion cases of childhood diarrhea worldwide each year, which, when they are not fatal, can cause enteric environmental dysfunction (EED), a subclinical condition characterized by intestinal inflammation and villous atrophy, and subsequently stunting (Budge et al., 2019; Syed et al., 2016). Although it is true that general transmission routes for diarrheal disease are known (e.g., fecal-oral contamination), the specific source of the pathogen (e.g., water or food) and the behaviors that are contributing to its spread vary by region, population, and context. Therefore, designing an effective intervention requires the precise identification of which behaviors need to be changed and under which circumstances. For example, in a study, which included 45 neighborhoods in 10 cities in 9 countries, researchers using direct measures were able to determine that infections were linked to exposure to contaminated water as well as the source of that water (e.g., open drains, flood water, and drinking water), which allowed for a more targeted intervention (Wang et al., 2022). Finally, direct measurement, even in other research areas in global health, can more easily capture event variation for each individual (e.g., when they wash their hands, how often they use soap and did it correctly) than indirect measures which typically can only capture overall variation across individuals.

The methods outlined in this paper might also be useful to researchers and practitioners who want to apply to behavioral observation and direct measurement of behavior in other areas of public health research and practice. Although the potential improvement in data quality with the use of direct measures has been demonstrated in other studies, detailed descriptions on how to collect those data are rarely included (e.g., Harvey, 2018; Teunis et al., 2016) For example, exclusive breastfeeding up until an infant reaches 6 months at which time complementary foods should be introduced is an essential component of both immediate and long-term growth outcomes and health. Using the methods outlined this paper, both researchers and practitioners would be able to record the frequency and duration of breastfeeding, the time that lapsed between feedings, environmental events that may cue feeding (e.g., infant crying), and any foods and the amount provided. The methods described could also be used to improve the quality of data on behaviors related to WASH interventions, such as latrine usage and handwashing. For example, with the use of subcategories, one could easily capture what was used to wash hands (e.g., water only, soap, sanitizer), how long the individual washed their hands for, as well as the events that proceeded and followed it (e.g., food preparation, feeding an infant).

Direct measures, even in smaller sample sizes, can also be used confirm the reliability of data collected through other methods and ensure implementation fidelity. Discrepancies in data gathered through indirect (self-report, interviews) and direct methods (observation) has been noted in previous research (e.g., Harvey, 2018). For example, during formative research when Harvey (2018) asked participants how often their chickens were out of their cage in a day, participants responded that the chickens were never out of their cages. However, when observations were conducted, enumerators noted that the chickens were out of their cages when being cleaned at least once per day. When asked about this, participants stated they assumed the researchers knew that the chickens would have to be out when cages were being cleaned. There are also circumstances when it is essential that an intervention or treatment is implemented with high levels of fidelity, in which case direct observation might be required. For example, healthcare providers use direct observation to ensure patients to take all doses of their tuberculosis medication (TB) to prevent multidrug resistant TB (i.e., Directly Observed Therapy Short Course; Yew, 1999). If looking to confirm events it is much easier to do so when the behavior of interest occurs frequently enough that it is likely to occur several times in the observation period (e.g., several times a day), occurs at a very specific time of day (e.g., Directly Observed Therapy), and reflects something the person is doing rather than what they are *not* doing (e.g., letting the chickens out of their cage). For example, it is easier to record a child being given and eating an egg (e.g., Baum et al., 2017) than it is to observe that a child has not being provided contaminated water or foods prior to 6 months.

### 5.2 Limitations

There are some limitations that need to be noted. The first is that compared to indirect measures, direct measures such as observation require additional privacy and ethical protections. Conducting observations in someone’s home, especially given the duration and the fact that these were women who had recently given birth can create an additional burden for study participants. We attempted mitigate this burden by hiring local women as enumerators, to increase the comfort level of the participants.

Another limitation is that although we attempted to maintain 80% agreement between data collectors for the duration of the study, we suspect that some observer drift, although minimal, did occur over the duration of the study, which indicator of less reliable data (see Figures 3 and 5). The physical distance between the trainers and enumerators, which was exacerbated due to travel restrictions related to the COVID-19 pandemic and political conflict in Ethiopia at the time, limited the type (e.g., in person) and amount of training that could be conducted. Although the trainers attempted to conduct booster sessions online over Zoom, unstable internet connections in Ethiopia made it difficult to turn on cameras and use videos for demonstration to provide immediate feedback. It is also possible that the duration of the observations themselves led to decreased attention on behalf of the enumerators and errors of omission. It is important to note that although in some cases highly accurate data is essential (e.g., tuberculosis treatment; Yew, 1999), it isn’t always required. For the purposes of this study slightly lower reliability of the data (inclusion of a greater or lesser number of events than what occurred) would not have negatively impact the quality of the results due to the total number of hours of observation and the high frequency of most events. occurring.

### 5.3 Conclusion

The purpose of this case study was to provide a comprehensive guide to conducting behavioral observation in epidemiological researchers and for public health professionals working in other areas both research and practice. Not only does it fill a gap in the literature on conducting behavioral observations to populate behavioral parameters (e.g., Weston et al., 2018), it also highlights how this type of data may be used to improve data quality in other key areas of global health such as WASH interventions. Although conducting behavioral observations is more resource heavy than self-report, the potential increase in data quality might be worth the expense given that it may lead to more precise and effective interventions, which will save resources in the long run. Future applications of this methodology may benefit from integrating real-time data quality checks, adapting definitions based on cultural context through participatory pre-testing, and replicating the protocol in diverse settings to enhance generalizability.

## Data Availability

We will post our data repository on GitHub. All data produced are available e upon reasonable request from the author.

https://github.com/YWAN446/EXCAM

